# Causal associations of body mass index and waist-to-hip ratio with acute appendicitis: a Mendelian randomization study

**DOI:** 10.64898/2026.03.11.26347691

**Authors:** Yingwen Qi, Youliang Wang, Tianwei Li, Sihui Gong, HongYan Zhao, Meiqin Jin, Wang Cuicui, Fan Li

## Abstract

**Background:** Acute appendicitis (AA) is a common surgical emergency. Observational studies have reported associations between obesity-related anthropometric traits and AA, but these associations may be affected by confounding and reverse causation. We used Mendelian randomization (MR) to investigate the potential causal effects of body mass index (BMI) and waist-to-hip ratio (WHR) on AA risk.

**Methods and Findings:** We obtained genome-wide association study (GWAS) summary statistics for BMI (ukb-b-19953), WHR (ieu-a-73), and AA (finn-b-K11_APPENDACUT) from the IEU Open GWAS database. We conducted single-variable MR (SVMR) and multivariable MR (MVMR) analyses. The primary estimator was inverse-variance weighted (IVW) MR, complemented by MR-Egger, weighted median, weighted mode, and simple mode methods. Instrument strength was assessed using the variance explained and F-statistics. Sensitivity analyses included Cochran’s Q for heterogeneity, MR-Egger intercept and MR-PRESSO global test for horizontal pleiotropy, leave-one-out analysis, and Steiger directionality testing. We mapped instrumental variants to cis-eQTL genes (eQTLGen) and performed GO and KEGG enrichment analyses.

In SVMR, genetically predicted BMI (OR 1.145, P = 0.0006) and WHR (OR 1.336, P = 0.0040) were associated with higher AA risk. Instruments were strong (BMI: 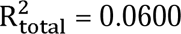; mean/min F-statistic= 64.15/29.76; WHR: 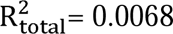; mean/min F-statistic= 48.41/29.75). Sensitivity analyses did not show strong evidence of heterogeneity or directional pleiotropy, and Steiger tests supported the hypothesized direction (exposure -> outcome). In MVMR including both traits, WHR remained associated with AA risk (OR 1.374, P = 0.0110), whereas BMI was not (P = 0.8000). Enrichment analyses suggested WHR-mapped genes were enriched in pathways related to adipocyte differentiation, while BMI-mapped genes were enriched in terms including nuclear envelope and endocytosis-related pathways.

**Conclusions:** These MR analyses are consistent with a potential causal relationship between obesity-related traits and AA risk, with WHR showing an association independent of BMI in multivariable models. Further work in diverse populations and with additional sensitivity analyses is warranted to assess robustness to pleiotropy and generalizability.

## Introduction

Acute appendicitis (AA) is a leading cause of acute abdominal pain requiring emergency surgery across the life course, with relatively higher incidence in adolescents and a modest male predominance [1]. The pathogenesis is multifactorial and may involve appendiceal luminal obstruction (e.g., lymphoid hyperplasia or fecalith), bacterial invasion, and contributions from environmental and neuro-immune mechanisms [2]. Although appendectomy is effective, perioperative morbidity and potential long-term sequelae highlight the need to identify modifiable risk factors that could inform prevention strategies [3].

Obesity, particularly central adiposity, has been linked to the incidence and severity of AA in observational studies. BMI is commonly used to quantify overall adiposity, whereas WHR captures fat distribution and central adiposity [5]. Prior studies have reported associations between higher BMI and more severe appendicitis and/or higher perforation risk [6], and imaging-based work suggests that obesity-related anatomical and vascular differences may influence diagnosis and inflammatory severity [7]. However, these findings may be influenced by residual confounding and reverse causation [4].

Mendelian randomization (MR) leverages germline genetic variants as instrumental variables to infer the causal effect of an exposure on an outcome, under key assumptions of relevance, independence, and exclusion restriction [8–10]. Because alleles are randomly allocated at conception, MR can reduce confounding and reverse causation compared with conventional observational analyses. Genetic evidence regarding the causal effects of BMI and WHR on AA risk remains limited. Therefore, we conducted both single-variable MR (SVMR) and multivariable MR (MVMR) analyses using publicly available GWAS summary statistics to assess the potential causal effects of BMI and WHR on AA, and to explore plausible biological pathways through functional annotation and enrichment analyses.

## Materials and Methods

### Data sources

GWAS summary statistics were obtained from the IEU Open GWAS database (https://gwas.mrcieu.ac.uk/): AA (finn-b-K11_APPENDACUT), BMI (ukb-b-19953; N = 461,460), and WHR (ieu-a-73; N = 212,244).

### Instrument selection and harmonization

For SVMR, SNPs associated with BMI or WHR at genome-wide significance (P < 5×10) were selected using extract_instruments in TwoSampleMR (v0.6.1) [11]. Linkage disequilibrium (LD) clumping was applied (r² = 0.001; window = 10,000 kb). Harmonization of alleles and effect directions was performed using harmonise_data.

For MVMR, SNPs associated with either exposure were obtained using mv_extract_exposures. Outcome data were extracted using extract_outcome_data with proxy searching enabled (r²>= 0.8). Harmonization used mv_harmonise_data, and covariate screening applied mv_lasso_feature_selection.

### Instrument strength and variance explained

To evaluate potential weak-instrument bias, we calculated the F-statistic for each SNP as *F_i_ = (β_x,i_/SE_x,i_)^2^*, where *β_x,i_* and *S_x,i_* are the SNP-exposure association estimate and its standard error. We summarized instrument strength using the mean (and minimum) F-statistic across all selected SNPs for each exposure.

The proportion of variance in the exposure explained by each SNP was approximated as 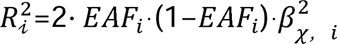, and the total variance explained was approximated by 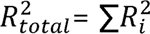 after LD clumping. The overall F-statistic for the instrument set was calculated as 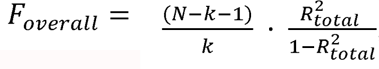, where N is the exposure GWAS sample size and k is the number of SNPs. We considered F-statistics > 10 as indicating sufficient instrument strength.

### MR assumptions

MR inference relies on: (1) IV relevance (genetic variants associated with exposure), (2) IV independence (variants not associated with confounders), and (3) exclusion restriction (variants affect the outcome only through the exposure).

### MR analyses

Primary causal estimates were obtained using IVW MR. Additional estimators included MR-Egger regression [12], weighted median [14], weighted mode [13], and simple mode [15]. Effect sizes are reported as ORs for AA per genetically predicted increase in BMI or WHR.

### Sensitivity analyses

We assessed heterogeneity using Cochran’s Q statistic (IVW) [16]. Horizontal pleiotropy was evaluated using the MR-Egger intercept and the MR-PRESSO global test (RSSobs and associated P-values). We conducted leave-one-out analyses to identify influential instruments and performed Steiger directionality tests to evaluate whether the instruments explained more variance in the exposure than in the outcome.

### Functional annotation and enrichment

cis-eQTL genes corresponding to instrumental SNPs were obtained from the eQTLGen database. GO and KEGG enrichment analyses were performed using clusterProfiler (v4.0.2) with significance threshold P < 0.05 [18].

### Ethics statement

This study used publicly available, de-identified GWAS summary statistics. No individual-level data were accessed, and no additional ethical approval or informed consent was required for this analysis.

## Results

### Single-variable MR: BMI and WHR as risk factors for AA

After instrument selection, 432 independent BMI-associated SNPs and 28 independent WHR-associated SNPs were retained. Instrument strength was adequate for both exposures (BMI: 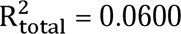; mean/min F-statistic= 64.15/29.76; WHR: 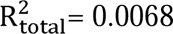; mean/min F-statistic= 48.41/29.75). In IVW analyses, genetically predicted BMI and WHR were associated with higher AA risk (BMI: OR 1.145, P = 0.0006; WHR: OR 1.336, P = 0.0040). Effect directions were generally consistent across complementary estimators (Table 1). Scatter, forest, and funnel plots supported an overall positive association (Figures 1A-1F).

**Table 1.** Results of MR analysis (IVW).

| Outcome | Exposure | Method | N SNPs | OR | P value |
| --- | --- | --- | --- | --- | --- |
| Acute appendicitis<br>(finn-b-K11_APPENDACUT) | BMI (ukb-b-19953) | IVW | 432 | 1.145 | 0.0006 |
| Acute appendicitis<br>(finn-b-K11_APPENDACUT) | WHR (ieu-a-73) | IVW | 28 | 1.336 | 0.0040 |

### Sensitivity and directionality analyses

We found no strong evidence of heterogeneity (BMI: Cochran’s Q P = 0.0600; WHR: P = 0.6100; Table 2). MR-Egger intercept tests did not indicate directional pleiotropy (BMI: P = 0.1855; WHR: P = 0.9668; Table 3). The MR-PRESSO global test did not provide evidence of horizontal pleiotropy (BMI: P = 0.061; WHR: P = 0.685; Table 4), although the BMI result was borderline and should be interpreted cautiously. Leave-one-out analyses did not identify single variants that materially drove the IVW estimates (Figures 2A-2B). Steiger tests supported the hypothesized direction from exposure to outcome for both BMI and WHR (Table 5).

**Table 2.**
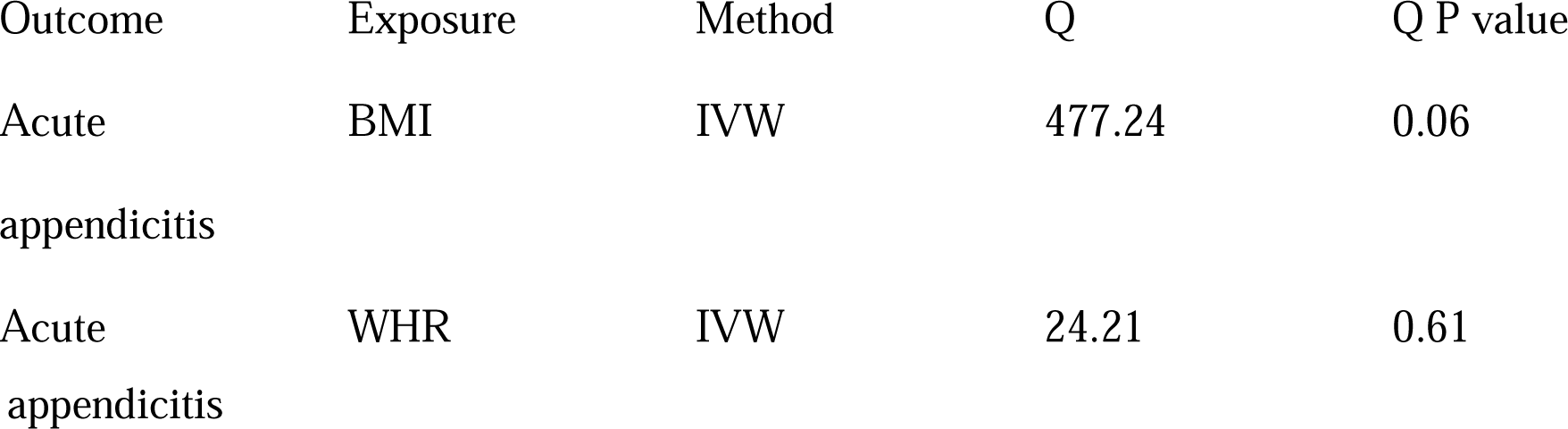
Heterogeneity test (Cochran’s Q).

**Table 3.**
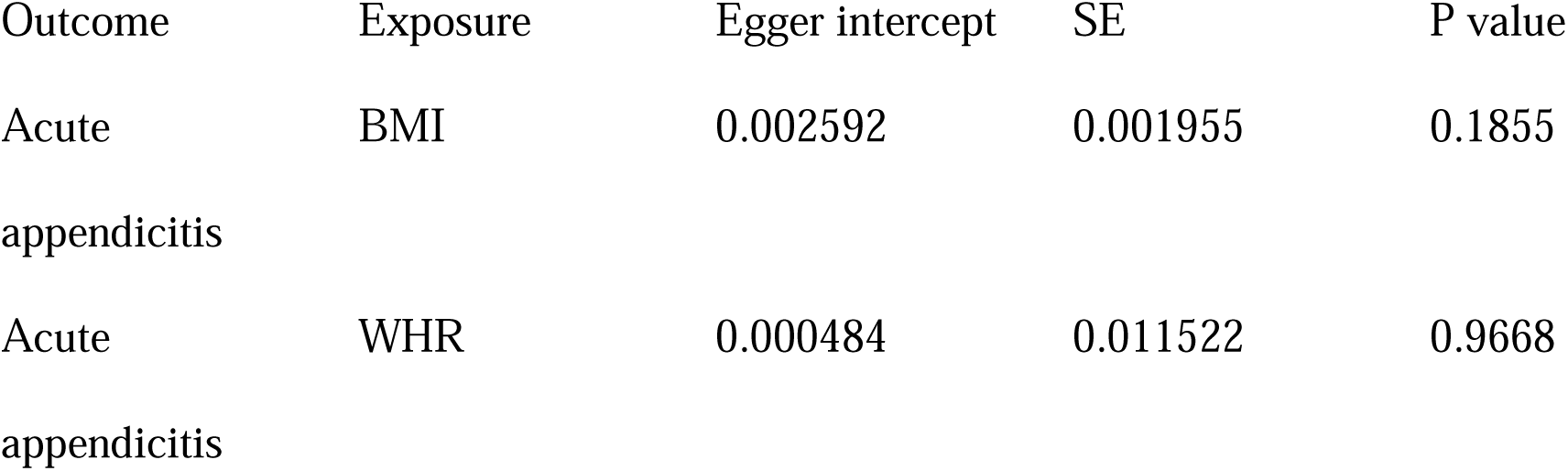
Pleiotropy test (MR-Egger intercept).

**Table 4.** Horizontal pleiotropy test (MR-PRESSO global test).

| Outcome | Exposure | RSSobs | P value |
| --- | --- | --- | --- |
| Acute appendicitis | BMI | 509.1532 | 0.061 |
| Acute appendicitis | WHR | 24.9990 | 0.685 |

**Table 5.** Steiger directionality test.

| Exposure | Outcome | SNPr<sup>2</sup>(exposure) | SNPr<sup>2</sup>(outcome) | Correct direction | Steiger P value |
| --- | --- | --- | --- | --- | --- |
| BMI | Acute<br>appendicitis | 0.061978773 | 0.002365553 | TRUE | 0 |
| WHR | Acute<br>appendicitis | 0.006739763 | 0.000152604 | TRUE | 4.35E-115 |

### Multivariable MR: WHR retains an independent effect

In MVMR analyses including both BMI and WHR, 316 BMI SNPs and 12 WHR SNPs were retained. WHR remained associated with AA risk (OR 1.374; 95% CI 1.076-1.757; P = 0.0110), whereas BMI was not (P = 0.8000; Table 6; Figure 2C).

**Table 6.**
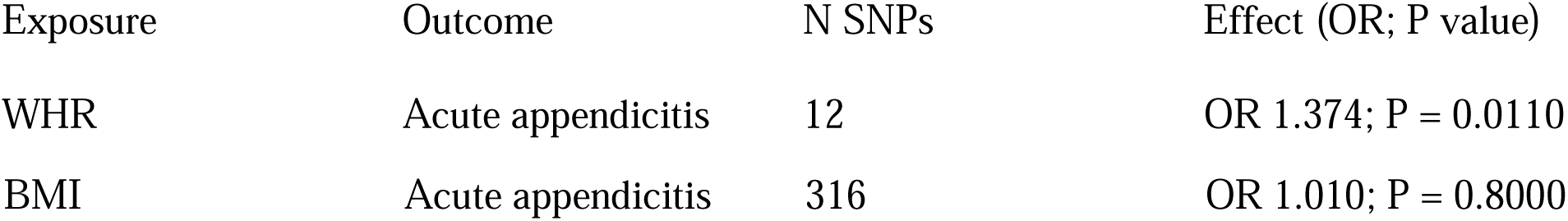
Multivariable MR analysis.

### Functional enrichment

BMI instruments mapped to 983 cis-eQTL genes, whereas WHR instruments mapped to 45 genes. BMI-mapped genes were enriched for GO terms including response to oxidative stress and nuclear envelope, and KEGG pathways including endocytosis and chemical carcinogenesis-reactive oxygen species. WHR-mapped genes were enriched for GO terms related to fat cell differentiation and KEGG pathways including non-alcoholic fatty liver disease and platelet activation (Figure 3A-3D).

## Discussion

Using MR, we observed associations consistent with potentially causal effects of BMI and WHR on AA risk in single-variable analyses. In multivariable models jointly accounting for both traits, WHR remained associated with AA risk whereas BMI did not, suggesting that central adiposity, as captured by WHR, may be more relevant to AA risk than overall adiposity.

Several mechanisms could plausibly link central adiposity to AA susceptibility, including obesity-associated alterations in gut microbiota, intestinal permeability, and systemic inflammation, which may modulate immune responses and promote appendiceal inflammation [19,20]. Immune dysregulation, including altered T-cell subsets and increased cytokine activity, could further contribute to inflammatory severity [21]. The enrichment results highlight adipocyte differentiation and metabolic/inflammatory pathways for WHR-mapped genes, providing biological plausibility for these hypotheses.

Strengths of this study include the MR design that reduces confounding and reverse causation, the use of multiple MR estimators, comprehensive sensitivity analyses, and MVMR to assess the independent effects of BMI and WHR.

Limitations should be considered. First, the GWAS datasets primarily included individuals of European ancestry, which may limit generalizability to other populations. Second, although MR-Egger and MR-PRESSO tests did not indicate strong pleiotropy, these methods have limited power in some settings and cannot fully exclude residual pleiotropy; the BMI MR-PRESSO result was borderline and warrants cautious interpretation. Third, the use of summary statistics precluded stratified analyses (e.g., pediatric-only AA) and limited exploration of non-linear or sex-specific effects. Fourth, UK Biobank and FinnGen are independent cohorts with distinct recruitment settings and population structure; therefore, the risk of direct sample overlap is likely minimal, but we cannot completely exclude overlap or relatedness without individual-level data. Future studies should validate these findings in non-European ancestry datasets, evaluate additional sensitivity analyses (e.g., radial MR or PhenoScanner-based confounder screening), and explore potential non-linear or subgroup-specific effects.

### Conclusions

Our findings are consistent with a potential causal relationship between obesity-related traits and AA risk. WHR showed an association that persisted after accounting for BMI in multivariable models, suggesting that central adiposity may play an important role in AA susceptibility. Further studies are needed to confirm generalizability and to strengthen inference regarding pleiotropy and mechanistic pathways.

## Data Availability

All GWAS summary statistics used in this study are publicly available from the IEU Open GWAS database (outcome: finn-b-K11_APPENDACUT; exposures: ukb-b-19953 and ieu-a-73).

## Funding

This study was supported by the Gansu Provincial Department of Science and Technology project “The Role of WHR and BMI in Lymphocyte and Cytokine Distribution in the Pathogenesis of Pediatric Appendicitis” and by the Gansu Provincial Natural Science Foundation (Grant No. 25JRRA331). The funders had no role in study design, data collection and analysis, decision to publish, or preparation of the manuscript.

## Competing Interests

The authors declare that they have no competing interests.

## Author Contributions (PLOS taxonomy-style)

Conceptualization: Fan Li

Methodology: Yingwen Qi, Youliang Wang, Fan Li Software: Tianwei Li

Formal analysis: Yingwen Qi, Tianwei Li

Investigation: Sihui Gong, HongYan Zhao, Meiqin Jin

Writing - original draft: Yingwen Qi, Fan Li

Writing - review & editing: All authors

Supervision: Fan Li

Funding acquisition: Fan Li

---

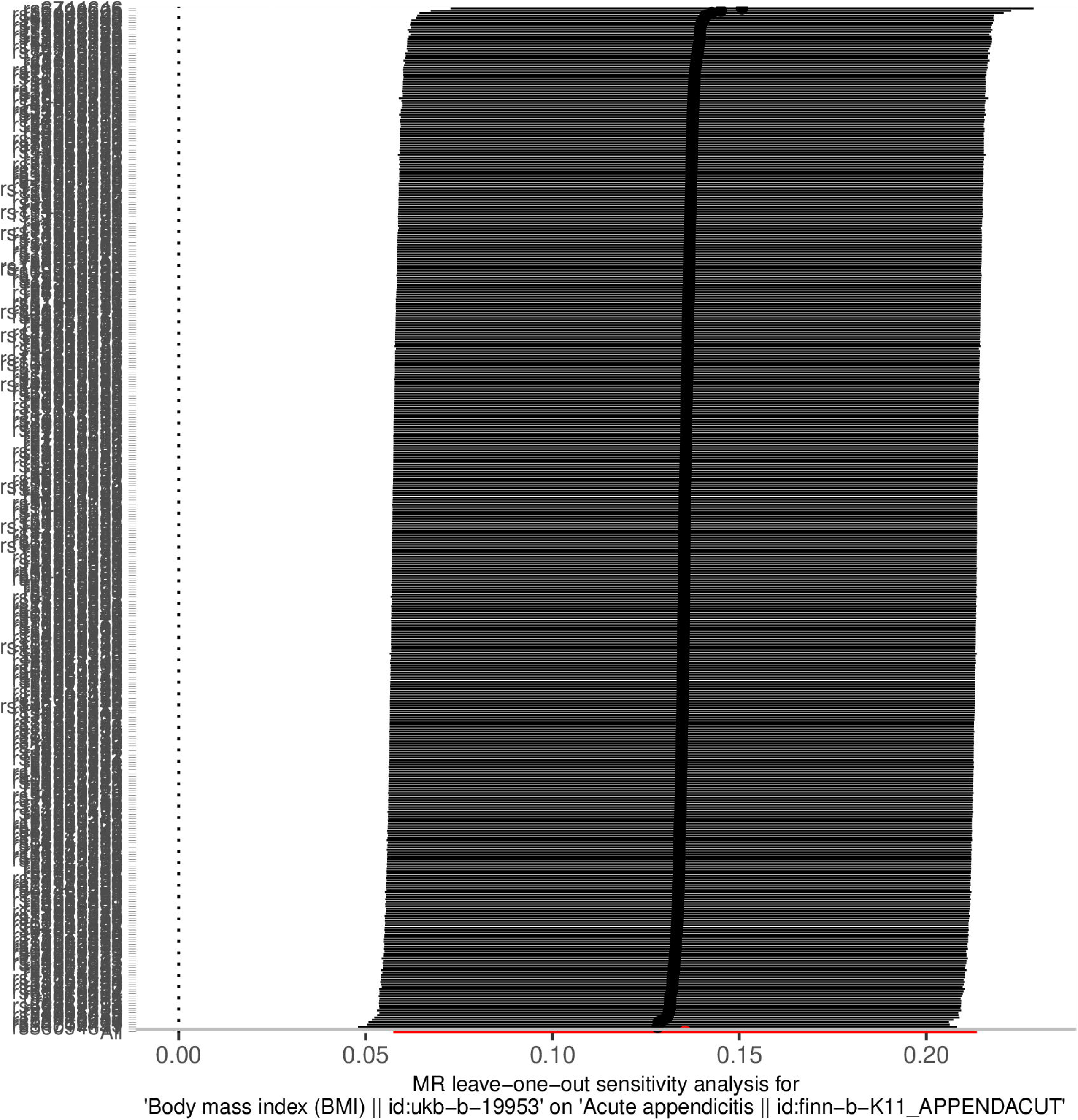

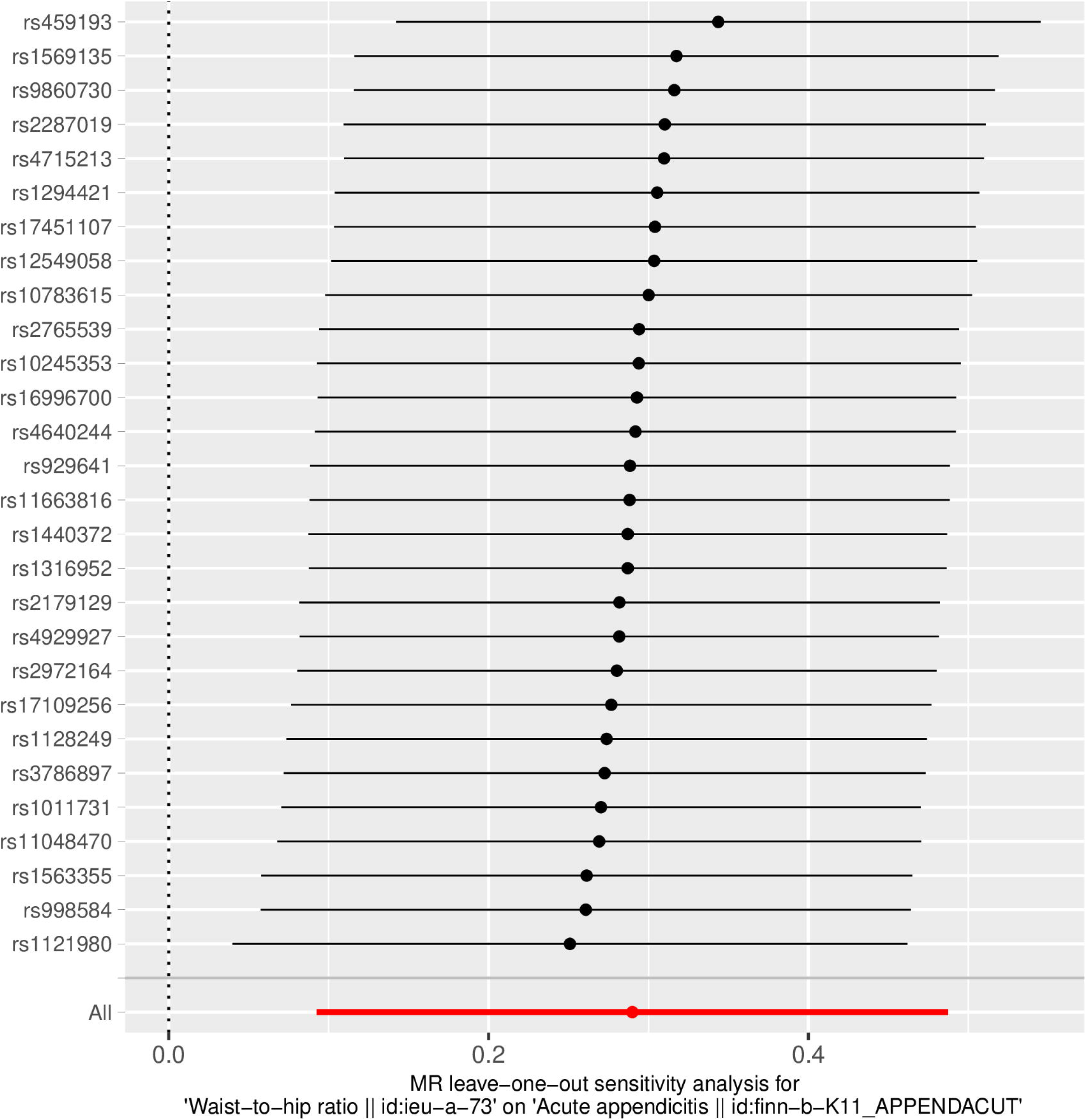

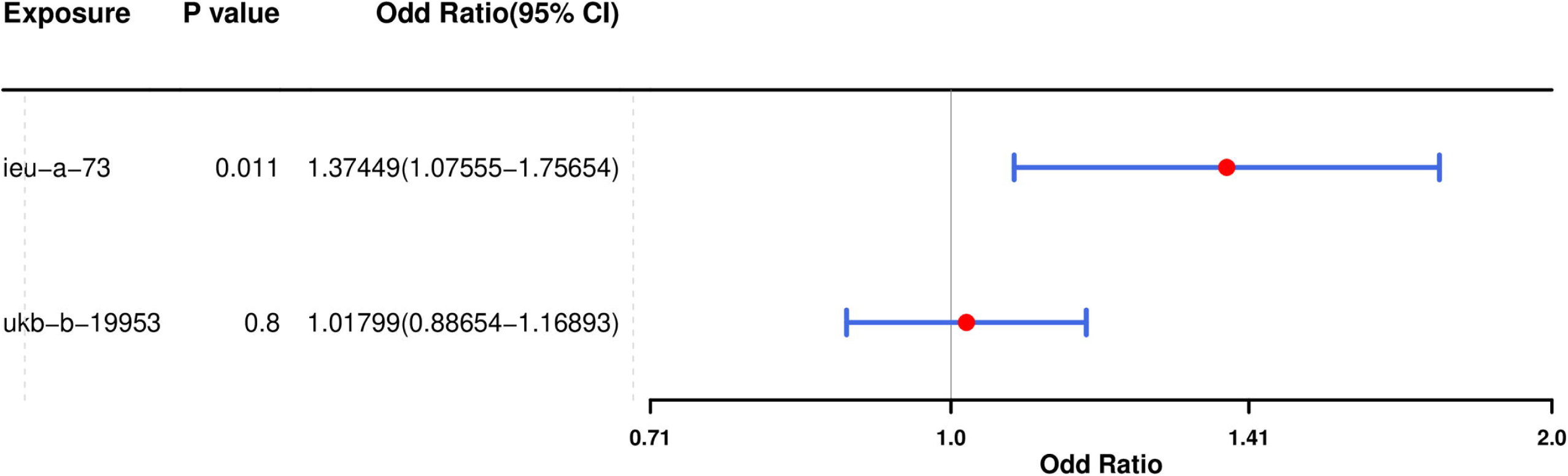

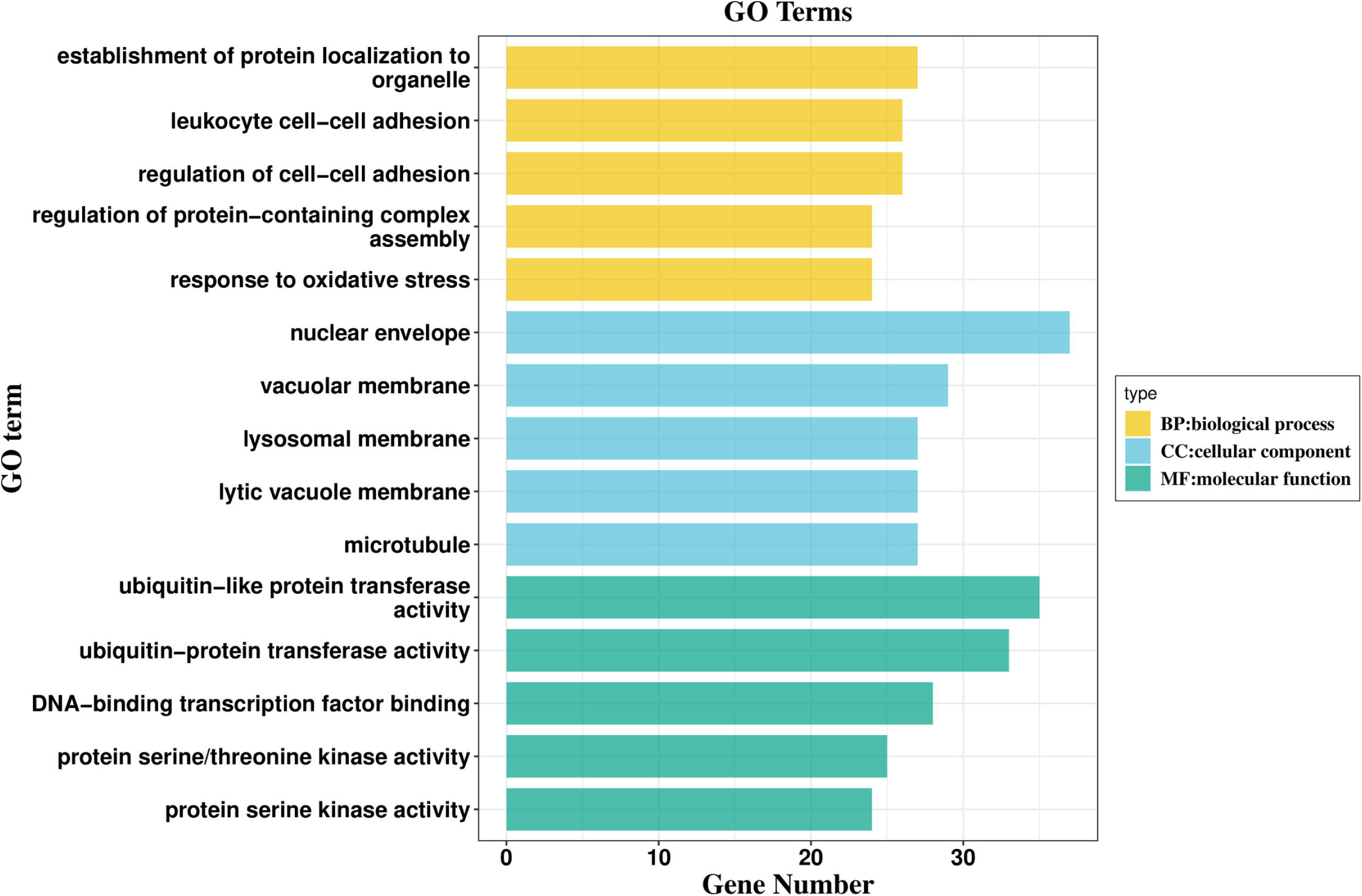

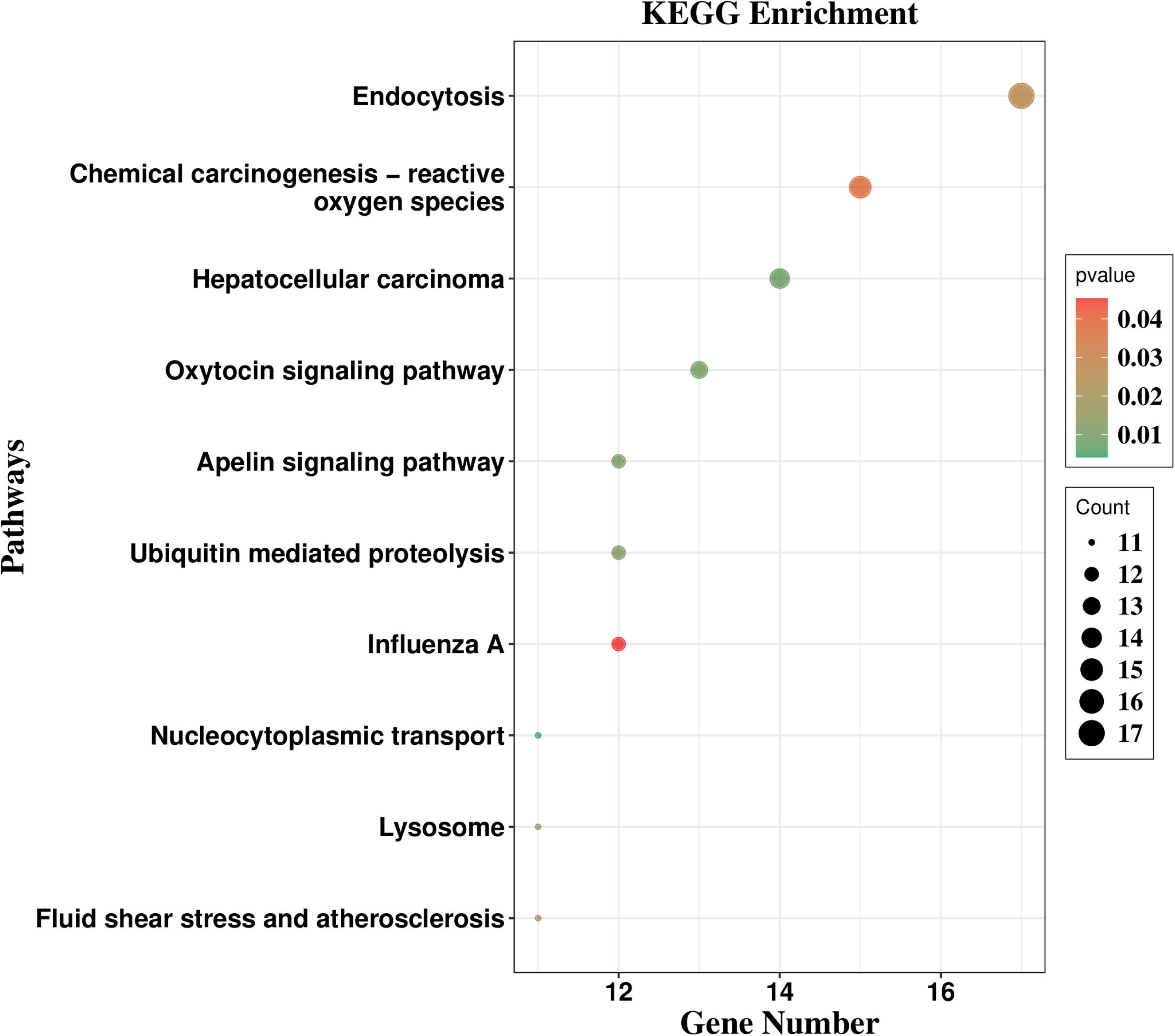

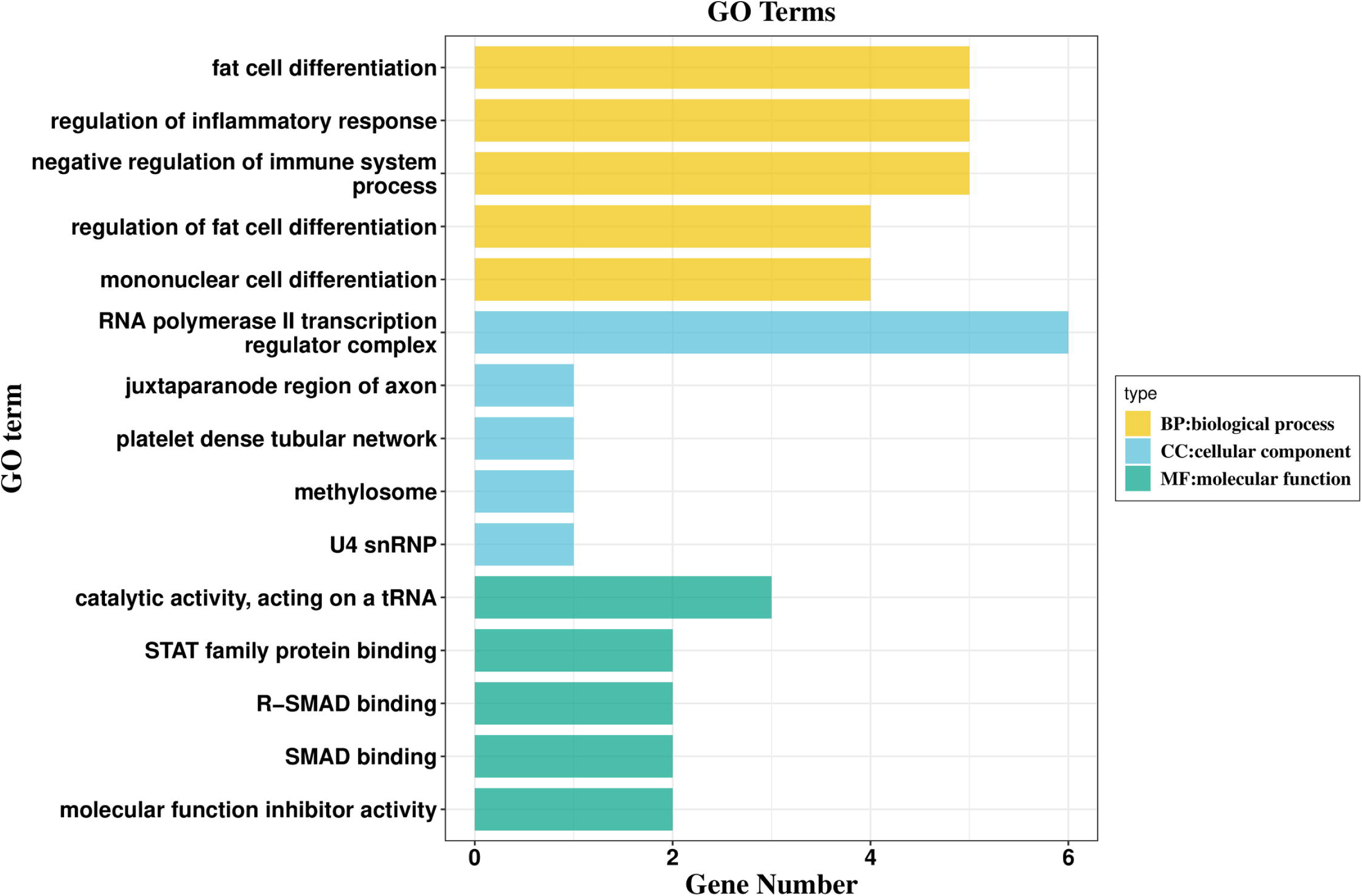

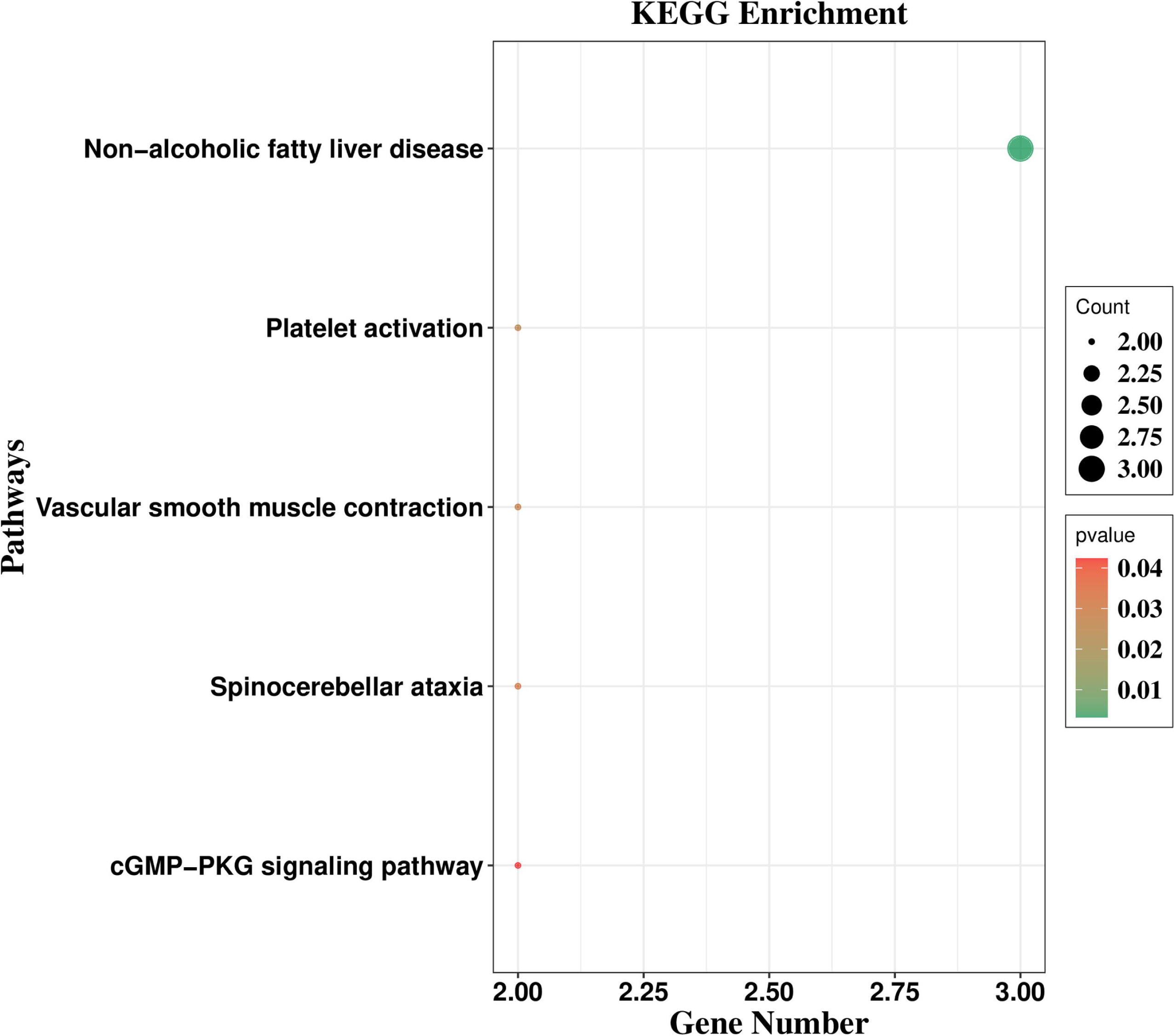

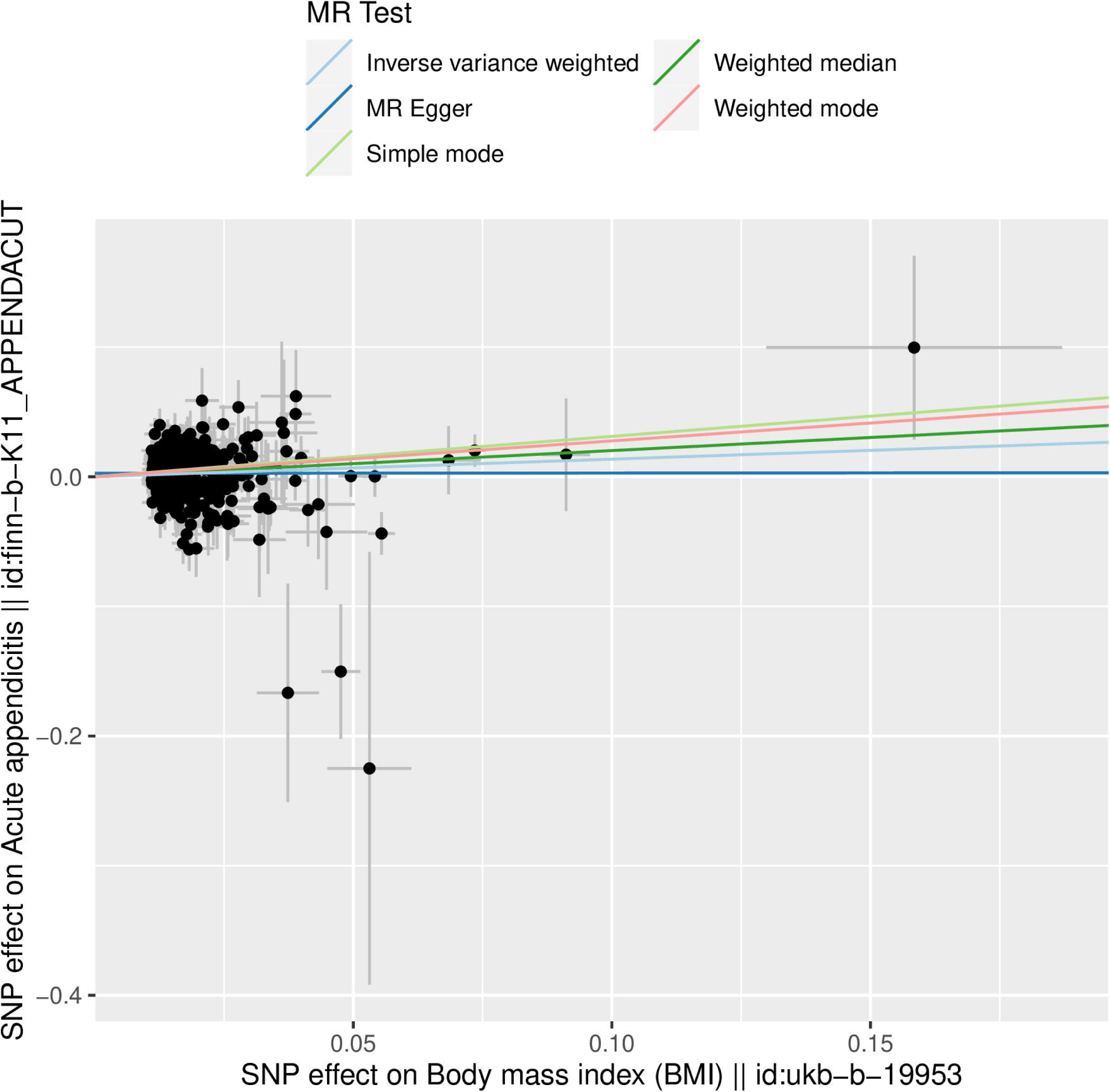

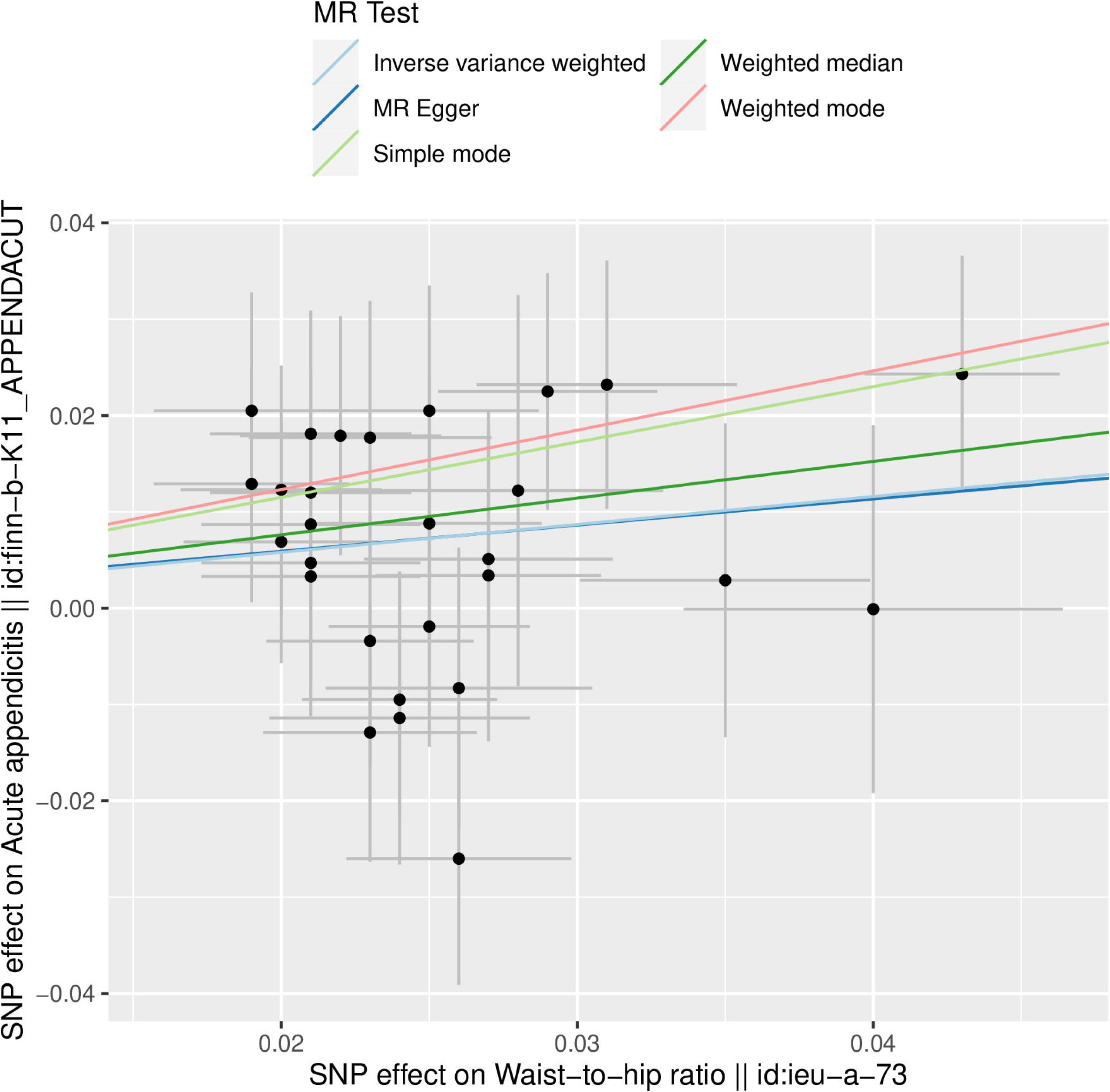

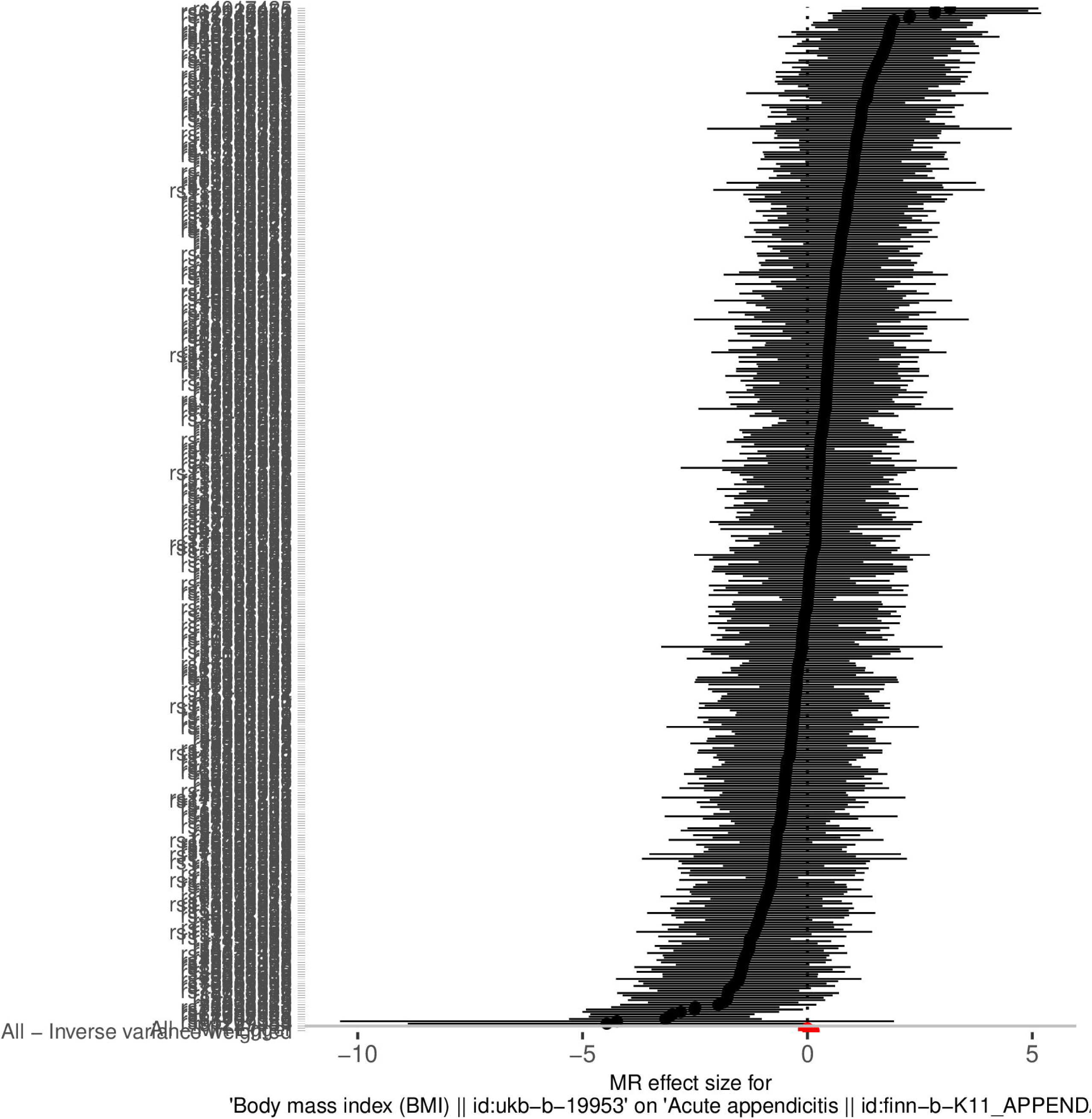

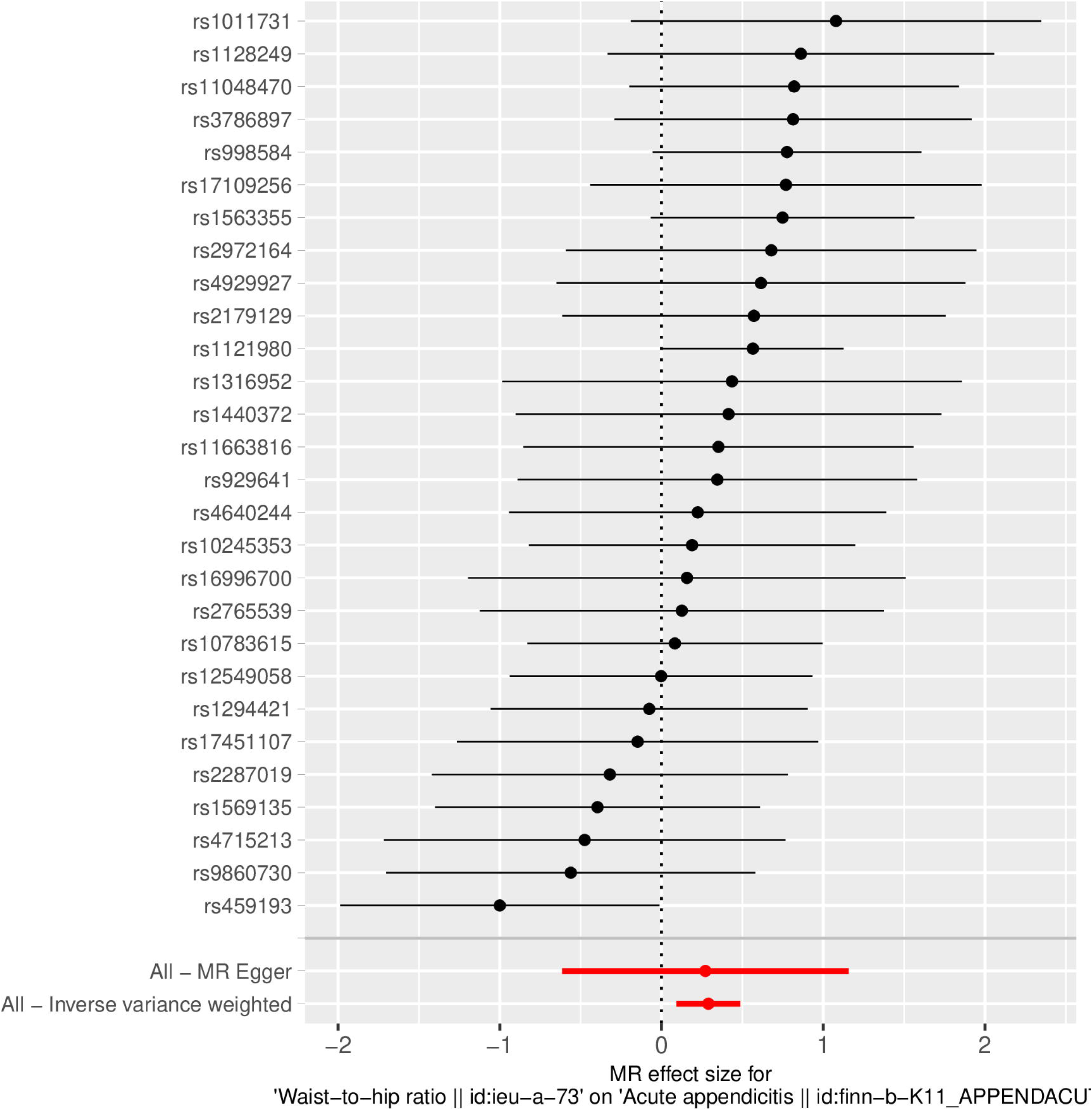

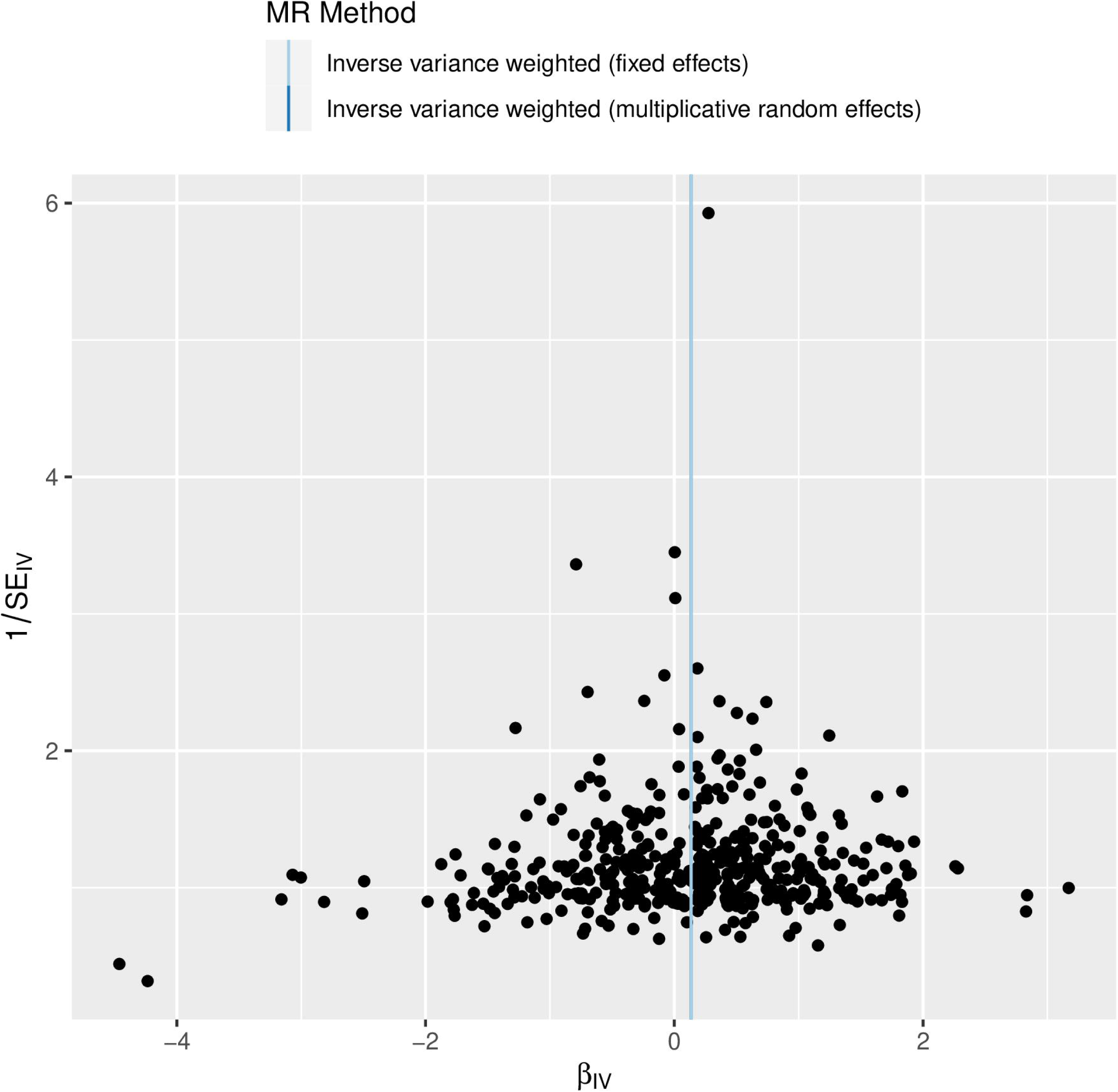

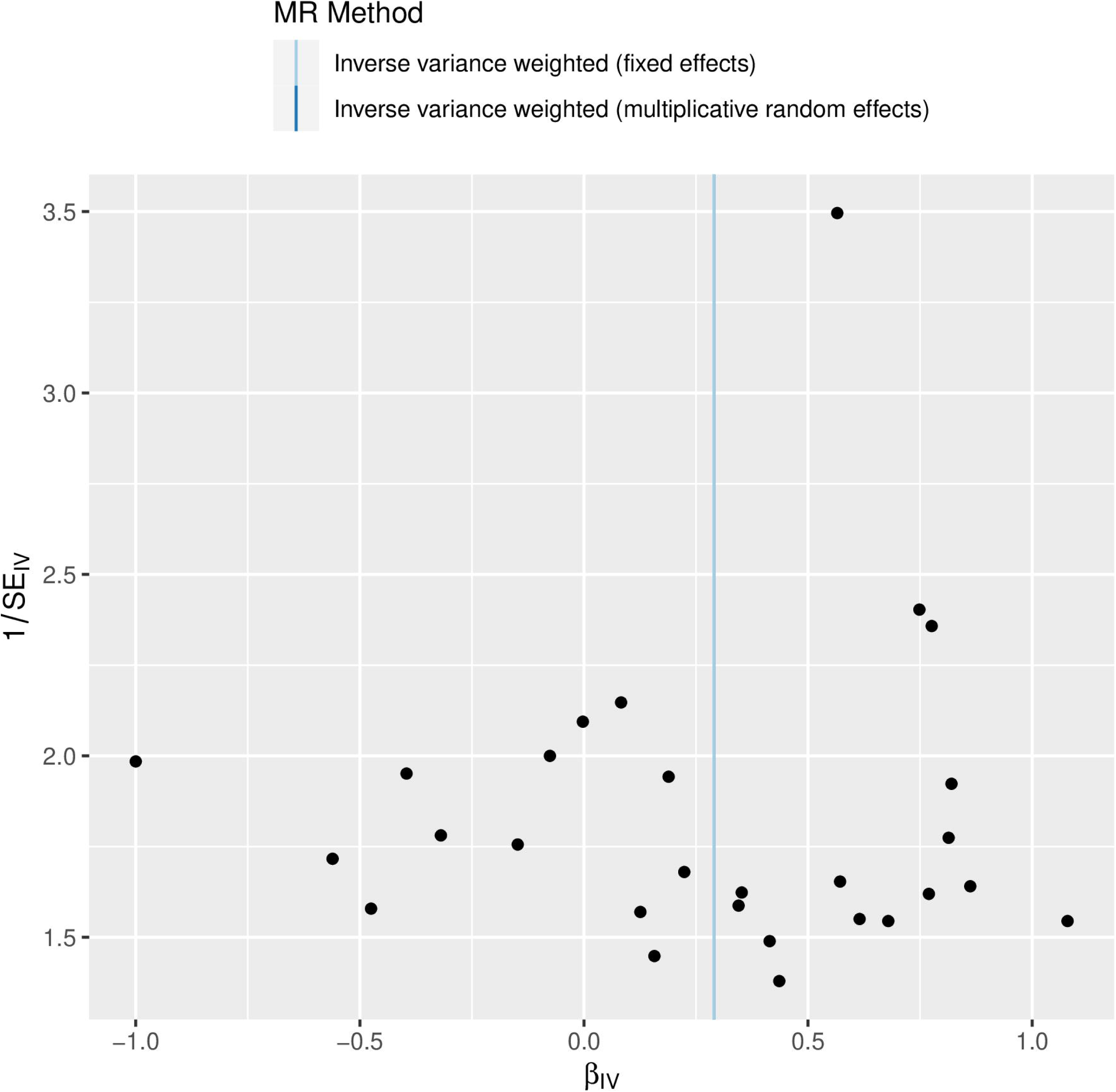

## Notes

### Competing Interest Statement

The authors have declared no competing interest.

### Funding Statement

This study was supported by the Gansu Provincial Department of Science and Technology project on the role of WHR and BMI in the distribution of lymphocytes and cytokines in the pathogenesis of pediatric appendicitis, and by the Gansu Provincial Natural Science Foundation (grant number 25JRRA331). The funders had no role in the study design, data collection and analysis, decision to publish, or preparation of the manuscript.

### Author Declarations

Ethics Committee of Gansu Provincial Maternal and Child Health Care Hospital waived ethical approval for this work. This study used only publicly available, de-identified summary-level GWAS data from the IEU OpenGWAS database and FinnGen. No new participants were recruited, and no individual-level identifiable data were used. Ethical approval and informed consent had been obtained in the original studies by their respective ethics committees.

